# Biological age acceleration measured by DunedinPACE associates most consistently with cognitive decline in elderly individuals

**DOI:** 10.64898/2026.03.23.26349074

**Authors:** Anna Marit Weißenburg, Marit Philine Junge, Jan Homann, Valerija Dobricic, Valentin Max Vetter, Ulman Lindenberger, Christina M. Lill, Ilja Demuth, Sandra Düzel, Lars Bertram

## Abstract

1

**Background:** Epigenetic clocks based on DNA methylation (DNAm) have emerged as promising biomarkers of biological aging, yet their associations with cognitive performance remain inconsistent. This study investigates the relationship between epigenetic age acceleration and cognitive performance in older adults using 14 DNAm clocks from five generations of development.

**Methods:** We analyzed data from the Berlin Aging Study II (BASE-II) using genome-wide DNAm profiles and cognitive assessments ascertained at baseline (T0) and two follow-up time points (T1, T2) in up to 1,014 individuals. DNAm-based age and age acceleration estimates were calculated using Biolearn and MethylCIPHER. Analyses focused on cross-sectional and longitudinal associations between DNAm clock estimates and cognitive performance, including sex-specific effects and comparisons with frailty as non-cognitive positive control.

**Results:** Among all tested DNAm clocks, DunedinPACE (a third-generation clock) showed the strongest and most consistent associations with cognitive performance. In addition, the fifth-generation SystemsAge framework also demonstrated robust associations with cross-sectional and longitudinal cognitive outcomes. In contrast, second-generation clocks (GrimAge [v2], PhenoAge) showed occasional nominal associations, while first-generation clocks (Horvath [v1], Hannum) and the causally-informed, fourth-generation clocks (e.g. YingCausAge, YingDamAge) showed no noteworthy signals. Likewise, telomere length estimated from DNAm was not strongly associated with cognitive performance in this dataset.

**Conclusions:** Our findings highlight DunedinPACE as a particularly informative biomarker for various aspects of cognitive aging, while other DNAm aging measures showed no consistent associations. Future work should further refine domain-specific epigenetic biomarkers to improve biological aging assessments and achieve a more reliable early detection of cognitive decline.

## 2 Background

Life expectancy has increased dramatically over the past century – from 46 years in 1948 to 73 years in 2023 – mainly owing to advances in medicine, sanitation, vaccination, antibiotics, and socioeconomic development [1]. However, increased longevity is accompanied by new challenges, such as age-related diseases and cognitive decline, as aging is associated with a loss of physical and cognitive function and, as a result, an increase in disease risk [2]. Aging is a heterogeneous process that varies substantially between individuals. This variability can at least partially be captured by estimating the “biological age” of an individual, which reflects underlying molecular and physiological changes [3].

Epigenetic modifications, particularly DNA methylation (DNAm), have emerged as key biomarkers of biological aging [4]. DNAm refers to the addition of methyl groups to cytosine residues, particularly at CpG dinucleotides, and plays a crucial role in gene regulation, cellular identity, and genome stability [5–7]. Changes in DNAm accumulate with age and have been linked to hallmarks of aging such as loss of proteostasis, mitochondrial dysfunction, stem cell exhaustion, and immune-senescence [8,9].

Leveraging this biological signal, epigenetic clocks have been developed to estimate biological age from DNAm patterns. First-generation clocks such as those by Hannum et al. (2013) and Horvath (2013) were designed to predict chronological age, and the difference between DNAm-based age and chronological age was proposed to represent an estimate of biological age [10,11]. Second-generation clocks, including PhenoAge and GrimAge were designed to enable a more direct estimation of biological age and disease vulnerability by incorporating health-related phenotypes and mortality risk at the stage of model building [12,13]. Third-generation DNAm clocks like DunedinPACE extend this approach by capturing the pace of biological aging using longitudinal biomarker and clinical data [14]. More recently, Ying et al. (2024) introduced a set of fourth-generation DNAm clocks (YingDamAge, YingCausAge, YingAdaptAge) representing CpGs selected based on their assumed causal effect(s) on aging, rather than solely estimating biological aging [16]. However, all these prior models share a common limitation: they yield a single, global estimate of biological age, thereby obscuring the fact that aging occurs at distinct rates across different physiological systems within the same individual. To address this, Sehgal et al. recently introduced the SystemsAge framework as a fifth-generation of DNAm clocks. By quantifying aging heterogeneity across specific functional domains – such as the inflammatory, hormonal, and metabolic systems – these models may provide a novel, multidimensional perspective on the biological aging process [17].

Epigenetic age acceleration (i.e., the deviation between DNAm-based age and chronological age) has been associated with numerous health outcomes. One limitation of many DNAm-based clock algorithms is that they were trained on DNAm profiles from blood samples. While blood is a commonly used tissue due to its relatively easy accessibility, it may only indirectly reflect aging processes in other organs, e.g. the brain [18,19]. This is partly the reason why the potential relationship between epigenetic aging and cognitive performance remains complex and has yielded largely inconsistent results across datasets. While some studies have reported significant associations between epigenetic age acceleration and lower cognitive performance or increased dementia risk [20–23], others did not observe such effects [24]. Discrepancies across studies may stem from the use of different clock algorithms, differences with respect to which cognitive domain was tested, reverse causation, sample size, or other cohort-specific factors [19].

In this study, we investigate the association between epigenetic age acceleration and cognitive function using multiple DNAm-based clocks from five generations of development. By doing so, we aim to clarify i) the extent to which epigenetic aging is linked to cognitive performance and decline, and ii) whether these relationships depend on the specific DNAm clock algorithm employed. Our analyses show clear and consistent associations between a range of cognitive performance measures and epigenetic age acceleration estimated by the DunedinPACE clock, while algorithms other development generations show only inconsistent and altogether weaker associations.

## 3 Materials and Methods

For an overview of the analyses performed in this study, please see Figure 1. All statistical analyses and data processing steps were performed using R version 4.4.0 (RRID:SCR_001905) and Python version 3.10.7 (RRID:SCR_008394).

**Figure 1:**
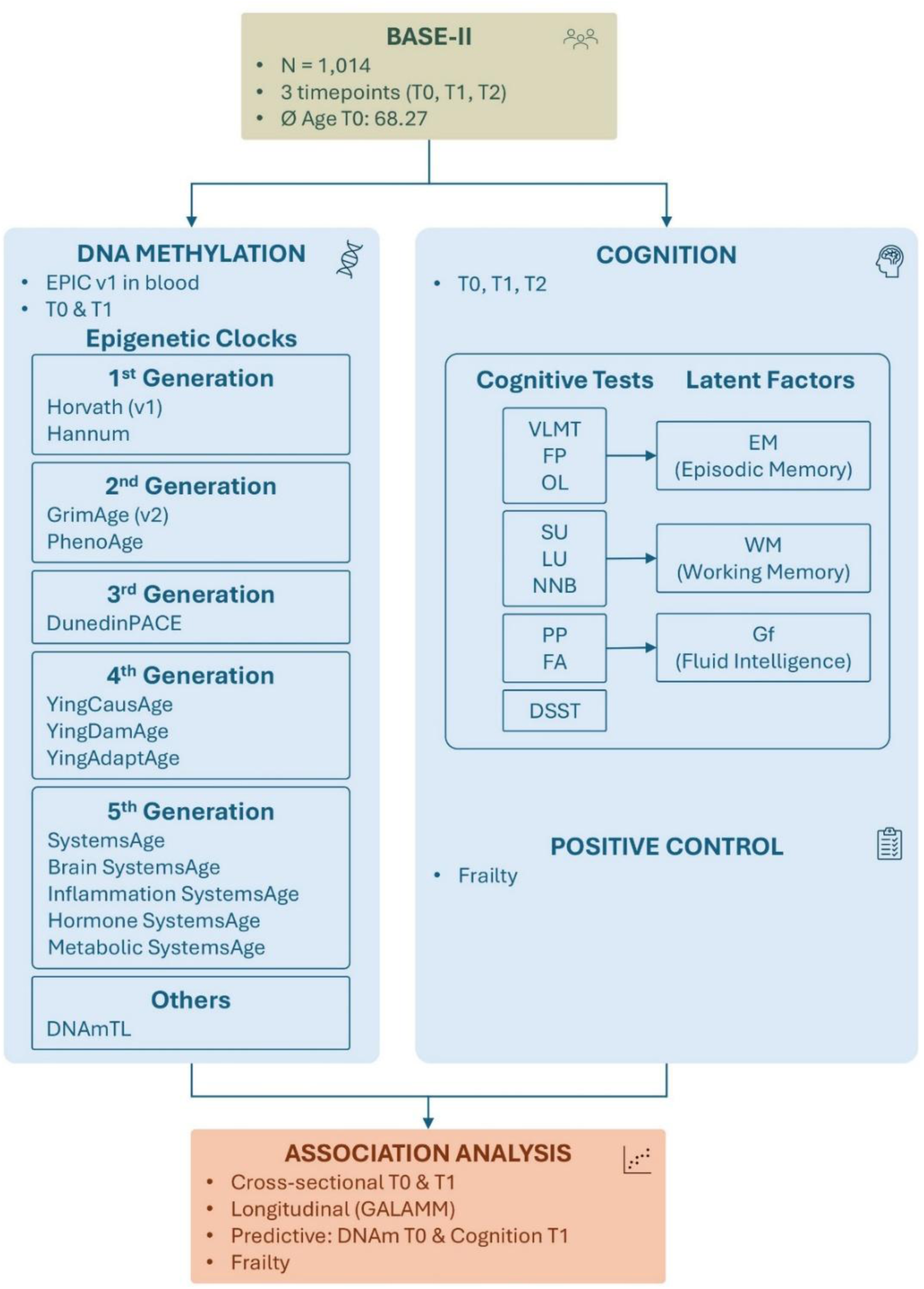
Schematic representation of the study design and workflow. Legend: Note that Gf and EM were constructed using additional cognitive tests which were not tested individually (Supp. Material).

### 3.1 Study Design and Participants

All analyses herein are based on data from the Berlin Aging Study II (BASE-II), a multidisciplinary cohort study designed to investigate healthy aging in the context of demographic change. The full BASE-II dataset includes approximately 1,600 older adults (60-85 years at baseline) and 600 younger adults (20-35 years at baseline) from the Berlin metropolitan area [25] Medical, genetic, and epigenetic data were collected and/or generated at two BASE-II sites: Charité-Universitätsmedizin Berlin and University of Lübeck. For this study, two waves of DNAm data were used (T0: 2009-2014, T1: 2018-2020). Cognitive data were ascertained at the Max Planck Institute for Human Development (MPIHD). Here, we make use of three waves of cognitive data collection (T0: 2013-2015, T1: 2018-2020, T2: 2022-2023) [25–27]. Matched DNAm and cognitive data were available for up to 1,014 individuals, with varying values at different times and for different tests (Supp. Table S1).

Concordance in epigenetic vs. clinical sex assignments was verified using the SexEstimator algorithm [28]. Two individuals with karyotypes 47,XXY and 45,X0 were excluded from the analyses.

### 3.2 DNA Methylation Data

#### 3.2.1 DNA Extraction, Processing and DNA Profiling

Genomic DNA was extracted using the Plus XL Manual Kit (LGC) following the manufacturer’s instructions, and concentration and quality was assessed via NanoDrop ONE spectrophotometry (Thermo Fisher Scientific, Waltham, MA, USA) as described before [29].

DNA methylation was measured using the Illumina Infinium MethylationEPIC BeadChip array version 1 (Illumina, Inc., San Diego, CA, USA) at the Institute of Clinical Molecular Biology (IKMB, UKSH campus Kiel) as described before [30]. Bisulfite conversion was performed using the EZ DNA Methylation Kit (Zymo Research, Irvine, CA, USA).

#### 3.2.2 Data Processing and Quality Control

Beta values were calculated as [31]:

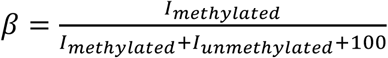

The DNAm data then underwent the following quality control (QC) assessments using the R package *bigmelon* [32]. Individuals were excluded if

i) the bisulfite conversion efficiency was below 80% according to the R function *bscon*,
ii) they had a detection p-value greater than 0.05 for more than 1% of the CpGs according to the *pfilter* function,
iii) they were outliers with respect to their *β* values according to the *outlyx* function,
iv) they showed a large change in *β* values after normalization using the R function *qual* (large change is defined as a deviation >0.1 of the root mean square of the *β* value (*rmsd*)).

Individual CpG probes were excluded from all subsequent analyses if

i) they had a detection p-value greater than 0.05 in more than 1% of individuals,
ii) they had a bead count smaller than 3 in 5% of individuals,
iii) they are assigned to positions that are influenced by SNPs [33],
iv) they are assigned to multiple positions according to Nordlund et al. [34],
v) they are located on the X or Y chromosomes,
vi) they are no CpGs.

### 3.3 Cognitive Function

The nine cognitive tests Digit Symbol Substitution Test (DSST), Figural Analogies (FA), Face Profession Task (FP), Letter Updating Task (LU), Number-N-Back Task (NNB), Object Location Task (OL), Practical Problem Task (PP), Spatial Updating Task (SU) and Verbal Learning and Memory Test (VLMT) were measured at the time points T0, T1, T2. Factor scores for episodic memory (EM), working memory (WM), and fluid intelligence (Gf) were derived using latent confirmatory factor analysis (Düzel et al., 2016). Some test results were removed due to outlier data (Supp. Material). Additional outliers (± 4 SD from the mean) in all cognitive tests were excluded. For better comparability, the cognitive test results were z-standardized. To estimate individual longitudinal changes in cognitive performance across the three available timepoints, we utilized Generalized Additive Latent and Mixed Models (GALAMMs). Specifically, we used the *galamm* R package, which implements the approach developed by Sørensen et al. (2023). This framework extends standard generalized linear mixed models by allowing for the simultaneous estimation of smooth population-level trajectories and subject-specific latent traits. Given the continuous nature of the cognitive scores, we specified a Gaussian GALAMM with an identity link function. The model decomposed the longitudinal trajectories into two components: a population-level mean trajectory estimated via penalized regression splines and subject-specific deviations. The latter were modeled as latent variables (random effects), comprising a random intercept for baseline performance and a random slope to quantify the individual rate of change over time. The individual longitudinal cognition score for each participant was defined as their estimated random slope coefficient. This value represents the subject-specific deviation from the average population change over the three timepoints [35].

### 3.4 DNA Methylation (DNAm) Clocks

As outlined in the introduction, existing DNAm clocks differ in the underlying models, functional focus, and training data. To evaluate and interpret differences with respect to each clock’s association with cognitive performance, we conducted a detailed comparison of various clocks from five different development generations. Specifically we selected two first-generation clocks – DNAmAgeHorvath (v1) [11] and DNAmAgeHannum [10] – as well as two second-generation clocks – DNAmAgePhenoAge [12] and DNAmAgeGrimAge (v2) [36]. From the third development generation we used the only available clock: DunedinPACE [14]. Furthermore, we used three recently developed fourth-generation DNAm clocks – YingCausAge, YingDamAge, and YingAdaptAge [15], which are based on a potential causal role of selected CpGs in aging. Additionally, we incorporated five models from the SystemsAge framework [17] to measure system-specific biological aging. Specifically, we selected general SystemsAge, along with the Inflammation, Hormone, and Metabolic SystemsAge clocks, owing to their associations with cognition and dementia in the Sehgal et al. (2025) study. We also included the Brain SystemsAge clock given its organ-specific relevance. Lastly, we also used telomere length (TL), whose shortening with age as a function of number of cell division is well established [37]. Here, TL was estimated using the DNAm-based estimate “DNAmTL” [38].

#### 3.4.1 Epigenetic Age Calculation

Epigenetic age estimates from each DNAm clock were derived using the open-source Python library *Biolearn* (version 0.6.5), which provides a standardized framework for the implementation and harmonization of both established and newly developed epigenetic clocks [39]. We calculated the predicted age using raw DNAm beta-values, chronological age (at the time of cognitive testing), and biological sex. Because the clocks from the SystemsAge framework are currently not included in *Biolearn*, their estimates were computed using the *methylCIPHER* (version 0.2.0) R package [40] using code provided in the primary SystemsAge publication [17]. The epigenetic age estimates were successfully computed for 1,157 participants at baseline (T0) and 1,155 participants at follow-up (T1). A complete overview of the calculated epigenetic age estimates and age acceleration residuals across both time points is provided in Supp. Table S2. Each clock relies on a specific set of CpG sites; missing CpGs were imputed in *Biolearn* during the computation process. To validate the *Biolearn*-based age estimates, selected DNAm clocks were also computed using the DNAmAge Calculator from the Clock Foundation [11,41], a widely used web-based application. This revealed largely identical estimates, with slight differences for the Horvath (v1) algorithm, probably due to differences in DNAm data normalization. For more details see Supp. Material and Supp. Table S11.

#### 3.4.2 Age Acceleration, Aging Rate and DNAmTL Adjustment

To assess differences in biological aging, we computed measures of epigenetic *age acceleration,* represented by the residuals from linear regression analyses of DNAm age on chronological age [4]. This approach, referred to as *relative age acceleration,* controls for the high correlation between epigenetic and chronological age. Individuals whose age acceleration values deviated by more than four standard deviations from the mean were excluded in an iterative process until no outliers remained. No age acceleration residuals needed to be computed for DunedinPACE as this clock reflects the pace of biological aging rather than an epigenetic age estimate. Specifically, it quantifies the rate at which an individual is aging biologically, with a value of 1.0 indicating one year of physiological change per chronological year, and higher or lower values representing faster (=accelerated) or slower (=decelerated) aging, respectively [14].

A similar “age acceleration” approach was applied to the DNAmTL data. Specifically, we adjusted for age using a linear regression approach and defined the residuals as adjusted DNAmTL (DNAmTLadj) [38]. To facilitate comparability across clocks, the signs of the YingAdaptAge and DNAmTLadj values were inverted so that higher values consistently indicated accelerated biological aging.

### 3.5 Statistical Models

To account for potential confounders in the DNA methylation data unrelated to cognition, a principal component analysis (PCA) was conducted separately for each time point (T0 and T1). To this end, we defined a subset of uncorrelated CpGs by randomly selecting one CpG from each 100 kb genomic region as described before [29,30]. PCA was then performed on the resulting 25.892 CpGs (T0) using the R function *prcomp*. The number of principal components (PCs) to include as covariates was determined using scree plots. Accordingly, the first five PCs were included for T0 and the first four PCs for T1 in all subsequent analyses. To adjust for genetic ancestry, we used the first six PCs from a PCA on genome-wide SNP genotyping data available in the same BASE-II individuals as described before [29].

Associations between age acceleration, aging rates (for DunedinPACE), DNAmTLadj, and cognitive performance were examined using multiple linear regression models (for cognitive performance treated as z-standardized continuous measures). Irrespective of cognitive status, we calculated pairwise inter-clock correlations using Pearson’s correlation coefficients for DNAm age calculated for all 14 clocks at T0.

All p-values were corrected for multiple comparisons using the Benjamini-Hochberg false discovery rate (FDR) at 5% [42].

#### 3.5.1 Linear Regression Models

Linear regression models were used to examine associations between age acceleration / aging rate / DNAmTLadj and cognitive test performance cross-sectionally at T0 and T1 (Equation 1) and longitudinally (GALAMM) (Equation 2) as well as a predictive model, i.e. testing T0 DNAm age estimates for an association with cognitive outcomes at T1 (Equation 3). Models were adjusted for sex, age at cognitive testing, DNA methylation PCs, and genetic PCs.

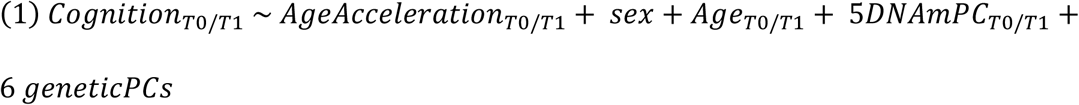

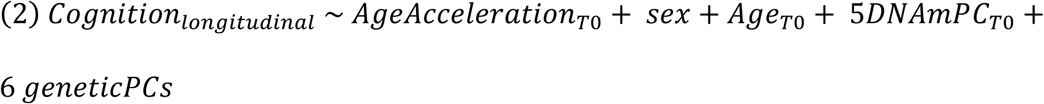

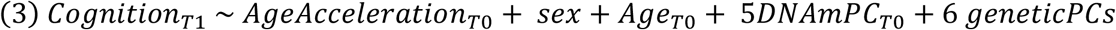

### 3.6 Frailty Index

To assess the biological validity of the general analytical pipeline used for the various cognitive outcomes, we used frailty as a “positive control“ since this (purely medical) trait showed relatively consistent associations with DNAm-based age accelerations in previous work [43,44], including BASE-II [45]. To this end, we used the “frailty index” as defined by Fried et al. (2001): here we categorized participants into non-frail (0 criteria), pre-frail (1-2), and frail (3-5) [46,47]. Frailty was assessed cross-sectionally at T0 and T1 (Equation 4), and longitudinally via the change in frailty index between time points (Equation 5). Cumulative logit models were applied to test for association between frailty and epigenetic age.

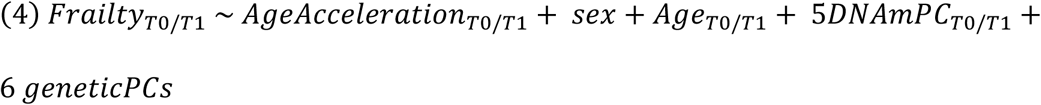

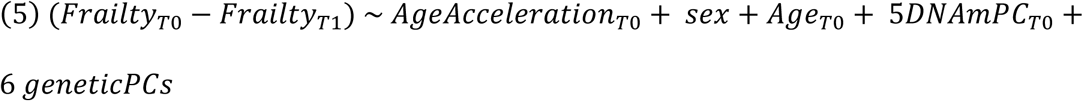

### 3.7 Sex-Moderated Interaction Analyses

Given prior evidence implying sex-specific differences in DNAm patterns [48,49], we conducted additional interaction analyses with sex as a moderator (Equation 7 and 8).

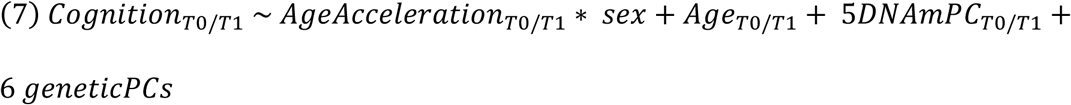

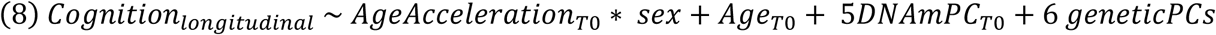

## 4 Results

### 4.1 DNA Methylation Clocks and Cognitive Function

#### 4.1.1 Age Acceleration and Inter-Clock Correlations

Overall, age acceleration, aging rate, and DNAmTL-adjusted values were predominantly weakly (r = 0.15–0.39) to moderately (r = 0.40–0.65) correlated across clocks (Figure 2). The strongest positive correlation was observed between the fourth-generation clocks YingDamAge and YingAdaptAge (r = 0.90), which both originate from the same general approach pinpointing causal CpGs. However, these fourth-generation clocks correlated only weakly (-0.12 ≤ r ≤ 0.19) with clocks from other development generations, with the exception of YingCausAge, showing a moderate correlation with the first-generation clocks (Hannum: r = 0.46, Horvath (v1): r = 0.47). We also observed moderate to strong positive inter-correlations, most notably between Brain SystemsAge and Metabolic SystemsAge (r = 0.78), as well as between general SystemsAge and Inflammation SystemsAge (r = 0.67). Interestingly, the SystemsAge models also demonstrated moderate positive correlations with several second- and third-generation clocks (e.g., general SystemsAge with GrimAge (v2): r = 0.62, and with DunedinPACE: r = 0.59), but showed negative correlations with YingAdaptAge (e.g., general SystemsAge: r = -0.64; Brain SystemsAge: r = -0.50). Among the earlier generations, the strongest correlation was observed between DunedinPACE and GrimAge (v2) (r = 0.64). Lastly, DNAmTL correlates most strongly with Hannum (r = 0.51) and Metabolic SystemsAge (r = 0.49), maintaining weak to moderate associations with the remaining clocks across all generations.

**Figure 2:**
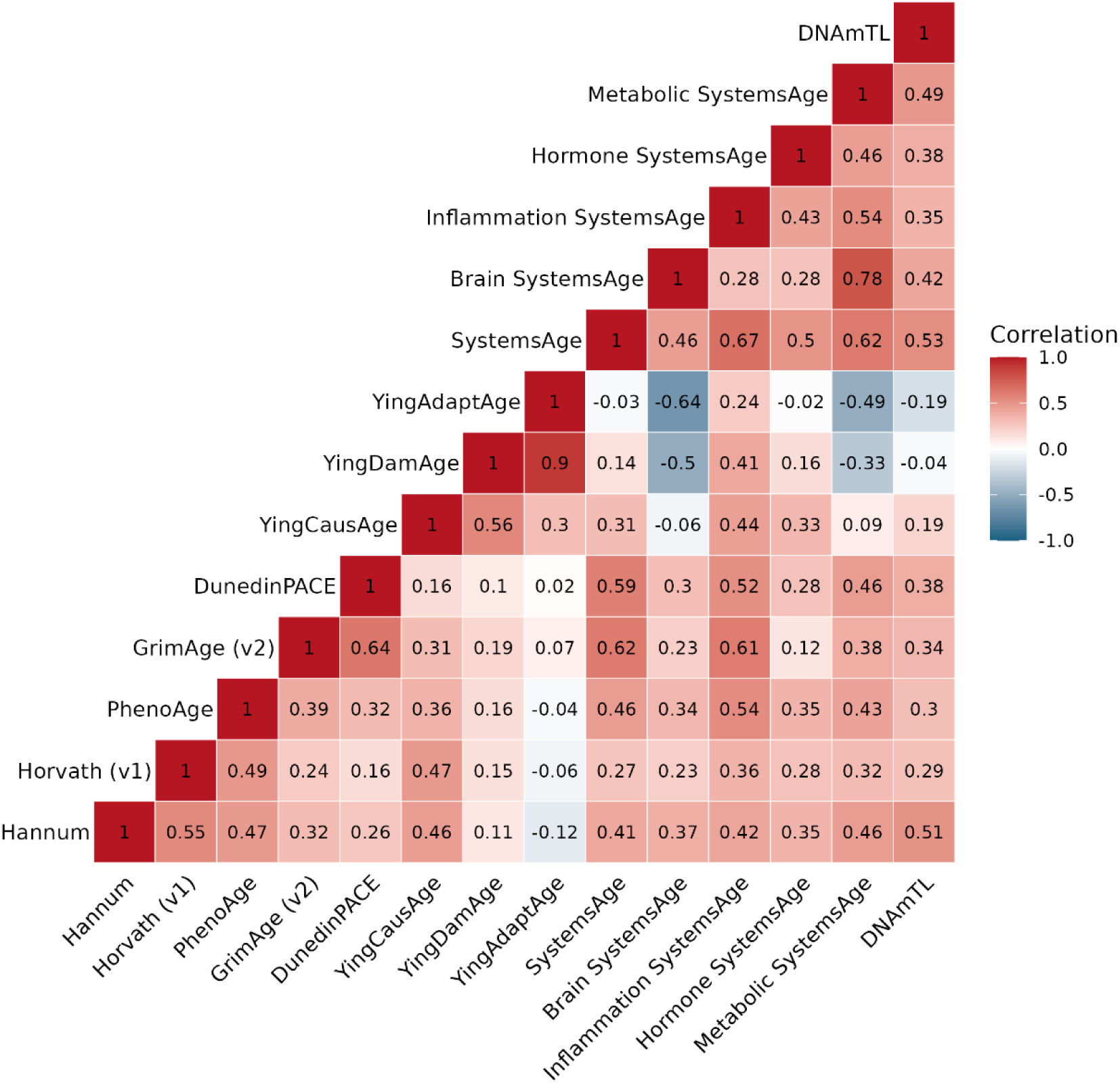
Correlations between DNAm clocks at T0. Legend: Pearson correlations between age acceleration, aging rate and DNAmTLadj of DNAm clocks. The age acceleration values of YingAdaptAge and DNAmTLadj were previously transformed by reversing the sign.

#### 4.1.2 Associations with Cross-Sectional Cognitive Performance

At T0, we identified 14 FDR-significant associations between cognitive test performance and different DNAm-based aging measures (Table 1). Particularly strong and consistent associations were observed with the DunedinPACE clock (i.e. 7 of 14 FDR-significant associations [Table 1] and 2 nominally significant associations [Supp. Table S3]). In other words, DunedinPACE showed at least nominal evidence of association with 9 of 12 tested cognitive traits. Notably, the recently introduced fifth-generation clocks also yielded robust individual effects: Inflammation SystemsAge yielded three FDR-significant associations, the general SystemsAge clock two, and the Hormone SystemsAge clock one.

**Table 1:** FDR-significant cross-sectional associations between DNAm clocks and cognitive performance at T0.

| DNAm clock | Cognitive test | N | P | P_adj | Beta | SE | R<sup>2</sup> |
| --- | --- | --- | --- | --- | --- | --- | --- |
| SystemsAge | FA | 938 | 1.85E-05 | 0.00218 | -0.0159 | 0.00370 | 0.0474 |
| DunedinPACE | FA | 940 | 2.59E-05 | 0.00218 | -1.337 | 0.316 | 0.0460 |
| DunedinPACE | WM | 1005 | 4.44E-04 | 0.0230 | -1.073 | 0.305 | 0.0500 |
| DunedinPACE | Gf | 1005 | 5.73E-04 | 0.0230 | -1.051 | 0.304 | 0.0518 |
| GrimAge (v2) | FA | 938 | 6.86E-04 | 0.0230 | -0.0364 | 0.0107 | 0.0400 |
| DunedinPACE | LU | 990 | 8.26E-04 | 0.0231 | -1.049 | 0.317 | 0.0188 |
| Hormone SystemsAge | LU | 989 | 0.00123 | 0.0285 | -0.0203 | 0.00626 | 0.0178 |
| SystemsAge | Gf | 1003 | 0.00136 | 0.0285 | -0.0114 | 0.00354 | 0.0513 |
| Inflammation SystemsAge | Gf | 1004 | 0.00169 | 0.0287 | -0.0144 | 0.00459 | 0.0505 |
| DunedinPACE | SU | 960 | 0.00171 | 0.0287 | -0.970 | 0.309 | 0.0694 |
| Inflammation SystemsAge | FA | 940 | 0.00222 | 0.0338 | -0.0147 | 0.00478 | 0.0376 |
| DunedinPACE | VLMT | 1001 | 0.00320 | 0.0449 | -0.917 | 0.310 | 0.0243 |
| Inflammation SystemsAge | WM | 1004 | 0.00358 | 0.0463 | -0.0134 | 0.004593 | 0.0471 |
| DunedinPACE | DSST | 938 | 0.00415 | 0.0498 | -0.910 | 0.317 | 0.0447 |

Taken together as a broader framework, the SystemsAge models collectively showed associations (nominal or FDR-significant) across 8 distinct cognitive traits. Next in terms of consistency is GrimAge (v2) showing association with 4 of 12 cognitive traits. In this context, the associations with the “figural analogies” test (FA) are particularly noteworthy: not only is this cognitive trait showing the statistically most significant associations with two clocks (i.e. general SystemsAge [p = 1.85E-05, p_adj = 2.18E-03] and DunedinPACE [p = 2.59E-05, p_adj = 2.18E-03]), but it also shows the largest number of FDR-significant associations in the cross-sectional analyses, i.e. 4 out of 14 (Table 1). Functionally, FA is considered an indicator of logical and visuospatial reasoning and belongs to the overarching domain of general fluid intelligence (Gf), which shows three FDR-significant associations itself (Table 1). Previous research has shown that the aging-related decline in Gf is particularly steep, more so than for other cognitive domains [50,51]. This could at least partially explain why cognitive outcomes related to Gf show the strongest and most consistent associations with DNAm age acceleration here.

Expanding the assessment to all 40 at least nominally significant associations observed at T0, it is noteworthy that – with one exception – first-generation clocks are absent from these findings (Supp. Table S3). It is also noteworthy that exactly half (i.e. 20 out of 40) of all significant associations (FDR and nominal combined) were observed with one of the three latent factor scores, again with Gf showing the largest number of associations (8x), followed by WM (7x) and EM (5x). This is not unexpected as latent factors are designed to capture more broad and encompassing aspects of cognitive function, which, as a result, associate more robustly with biological age by minimizing task-specific variance and random measurement error. Lastly, the observed directions of effect are without exception consistent with the overall hypothesis, i.e. for all significant associations a better test performance is associated with a lower age acceleration/aging rate (indicated by negative beta coefficients).

Interestingly, we observed no FDR-significant associations between any of the tested cognitive variables (incl. latent factors) and DNAm-based age measures in cross-sectional analyses of data ascertained at T1. Although the 26 nominally significant associations (Supp. Table S4) clustered around DunedinPACE (10x), the SystemsAge framework (general SystemsAge (4x), Inflammation (4x), Hormone (2x), Metabolic (1x), Brain (1x)) and GrimAge (v2) (3x)), similar to the findings at T0. The vast majority (i.e. 11 out of 14) of the FDR-significant pairs from T0 also achieved at least nominal significance in T1, including all associations with DunedinPACE. In agreement with these qualitatively overlapping patterns of association, the beta coefficients of the nominally significant pairs in T0 also correlate very strongly with the beta values in T1 (r = 0.95) indicating a much higher correspondence of associations between T0 and T1 than expected by mere comparison of p-values.

The attenuation of statistical significance from baseline (T0) to follow-up (T1) likely reflects a shift in the primary drivers of cognitive variance rather than a true disappearance of the epigenetic signal. At an average age of 70, subclinical biological aging, as captured by the utilized DNAm clocks, accounts for a detectable share of cognitive differences. However, by age 78, emerging age-related neuropathologies, such as accumulating vascular burden or neurodegenerative protein deposits, may begin to dominate cognitive decline, effectively drowning the epigenetic signal. Furthermore, cognitive floor effects at advanced ages can compress the variance in test scores, while the DNA methylome itself may become increasingly disordered, reducing the dynamic range and predictive precision of epigenetic estimators. This combination of competing clinical pathologies, restricted cognitive variance, and DNAm clock saturation systematically shrinks the effect sizes, which disproportionately impacts statistical significance and, in particular, threshold-dependent multiple testing corrections like the FDR approach.

Interestingly, the Brain SystemsAge clock did not show statistically significant associations with any of the analyzed cognitive traits at either time point.

#### 4.1.3 Associations with Longitudinal Cognitive Performance

The analysis of longitudinal associations between epigenetic measures of aging and cognitive test performance, estimated using the GALAMM framework ([35] and Methods), also revealed 13 FDR-significant findings. Similar to what was observed for both cross-sectional time points, DunedinPACE and various SystemsAge clocks emerged as the most robust DNAm predictors. Specifically, age acceleration measured with DunedinPACE showed the strongest and most robust longitudinal associations with five different cognitive tests: FA (p = 9.84E-05, p_adj = 0.00747), DSST (p = 1.37E-04, p_adj = 0.00757), SU (p = 1.78E-04, p_adj = 0.00757), LU (p = 0.00170, p_adj = 0.0317) and NNB (p = 0.00201, p_adj = 0.0317; Table 2). The DNAm clock with the second most consistent associations were those calculated by the SystemsAge framework which accounted for seven of the FDR-significant associations. Individually, these effects were driven by Inflammation (4x), Hormone (2x), and the general SystemsAge clock (1x). The only other clock represented among the FDR-significant associations was GrimAge (v2) (with trait DSST; p = 0.00154, p_adj = 0.0317).

**Table 2:** FDR-significant associations between DNAm clocks and longitudinal cognitive performance.

| DNAm clock | Cognitive test | N | P | P_adj | Beta | SE | R<sup>2</sup> |
| --- | --- | --- | --- | --- | --- | --- | --- |
| DunedinPACE | FA | 995 | 9.84E-05 | 0.00747 | -1.209 | 0.309 | 0.0322 |
| DunedinPACE | DSST | 999 | 1.37E-04 | 0.00747 | -1.177 | 0.307 | 0.0410 |
| DunedinPACE | SU | 998 | 1.78E-04 | 0.00747 | -1.135 | 0.302 | 0.0710 |
| SystemsAge | FA | 993 | 7.26E-04 | 0.0229 | -0.0122 | 0.00360 | 0.0284 |
| GrimAge (v2) | DSST | 997 | 0.00154 | 0.0317 | -0.0328 | 0.0103 | 0.0357 |
| DunedinPACE | LU | 1004 | 0.0017 | 0.0317 | -0.978 | 0.311 | 0.0112 |
| Inflammation SystemsAge | FA | 994 | 0.00184 | 0.0317 | -0.0145 | 0.00465 | 0.0268 |
| DunedinPACE | NNB | 1004 | 0.00201 | 0.0317 | -0.949 | 0.306 | 0.0396 |
| Hormone SystemsAge | DSST | 998 | 0.00251 | 0.0351 | -0.0187 | 0.00616 | 0.0354 |
| Inflammation SystemsAge | DSST | 998 | 0.00335 | 0.0423 | -0.0136 | 0.00463 | 0.0347 |
| Inflammation SystemsAge | FP | 1003 | 0.00370 | 0.0424 | -0.0137 | 0.00472 | 0.00235 |
| Hormone SystemsAge | LU | 1003 | 0.00441 | 0.0451 | -0.0178 | 0.00622 | 0.00935 |
| Inflammation SystemsAge | PP | 1003 | 0.00465 | 0.0451 | -0.0132 | 0.00464 | 0.0334 |

In addition to these 13 FDR-significant associations, 18 nominally significant signals were identified, the majority of which were driven by one of the SystemsAge clocks: i.e. general SystemsAge (4x), Inflammation SystemsAge (3x), and Hormone SystemsAge (3x) models. In agreement with the cross-sectional analyses and overall expectation, we again observed negative beta coefficients for all at least nominally significant associations indicating that accelerated biological aging is associated with a steeper cognitive decline (Table 2 and Supp. Table S5).

The last analysis paradigm we ran was to “predict” cognitive performance at T1 from epigenetic measures at T0. While a robust pattern of 34 nominally significant associations emerged, none of these remained significance after multiple testing correction using FDR, likely for the same reasons already outlined for T1 above. Among the nominally significant signals, associations with DunedinPACE again showed the strongest statistical support and overall seven significant associations (Supp. Table S5). As for the other analysis paradigms presented above, clocks from the SystemsAge framework represented the runners-up altogether accounting for 20 of the 34 nominal associations. Individually, these effects were driven by Inflammation (8x), general SystemsAge (7x), and Hormone (5x). Some additional nominally significant associations were observed for second-generation clocks (GrimAge (v2) and PhenoAge) and DNAmTLadj (4), while first-generation and fourth-generation clocks showed no significant predictive value (for full analysis see Supp. Table S6).

Similar to our observations in the cross-sectional analyses, the Brain SystemsAge clock again did not show statistically significant associations with any of the tested cognitive traits in the two longitudinal paradigms.

#### 4.1.4 Sex-Moderated Interaction Analyses

Given the established sex differences in aging – both on the DNAm level as well as cognitive performance [48,49] – we investigated whether any of the observed associations between DNAm clocks and cognitive performance differed by biological sex. This was tested using interaction analyses with biological sex as a moderator variable. These analyses did not reveal any FDR-significant evidence for an interaction effect moderated by this variable, and showed only few nominal significant interactions (Supp. Table S7-S9). Collectively, these results suggest that there is no noteworthy difference with respect to sex in the associations between DNAm-based aging acceleration and cognitive performance, at least not in the dataset analyzed here.

### 4.2 DNA Methylation Clocks and Frailty

As expected and previously reported in several studies [43–45], various FDR-significant associations were found between DNAm-based age acceleration and the frailty index. Similar to the cognitive outcomes, DunedinPACE showed the strongest and most consistent associations with frailty at both time points. With a p-value of 1.8×10-7 (p_adj = 2.5×10-6), the association between DunedinPACE and frailty at T1 showed, by a margin, the strongest support of all comparisons tested in this study. At the same time point, DunedinPACE was joined by robust, FDR-significant associations from GrimAge (v2) and four clocks from the SystemsAge framework (Inflammation, general, Hormone, and Metabolic SystemsAge) (Supp. Table S10a-b). Furthermore, nominally significant associations with longitudinal changes in frailty were observed not only for DunedinPACE, but also for the Inflammation and Metabolic SystemsAge clocks. In contrast, none of the other tested clocks, including all first- and fourth-generation clocks, showed more than nominally significant associations with this non-cognitive trait (Supp. Table S10).

## 5 Discussion

This study aimed to investigate the associations between DNAm-based biomarkers of aging and cognitive performance in a dataset of elderly adults from Germany. By examining 14 DNAm clocks from all development generations, we evaluated whether composite measures of age-related DNAm patterns were associated with a range of cross-sectional and longitudinal cognitive outcomes. To the best of our knowledge, our study is the first to systematically test clocks from the recently reported SystemsAge framework with respect to cognitive performance beyond the original report. Across all analysis arms, DunedinPACE emerged as the most consistent, and in all but one case, also most significant predictor of cognitive performance followed by one or more SystemsAge clocks. In contrast, the remaining clocks, including the fourth-generation approaches based on causal CpGs, showed only weak, inconsistent, or no associations. These findings are in line with prior work suggesting that DunedinPACE is the most powerful algorithm to capture biological aging and aging rates across a range of domains [14,22,23,52].

Currently available DNAm clocks differ in their underlying designs, which helps explain the heterogeneity of the observed associations with cognitive performance. Specifically, DNAm clocks of the first generation, such as Horvath (v1) and Hannum, were developed to estimate chronological age rather than functional aging processes. As expected, and consistent with previous studies [53,54], these clocks showed only weak and predominantly non-significant associations with the cognitive traits tested here.

Second-generation clocks, including PhenoAge and GrimAge, were trained on morbidity- and mortality-related outcomes and therefore capture a broader range of the biological aging process. Compared to PhenoAge, GrimAge (v2) showed the clearest and most consistent pattern, including two FDR-significant and several nominal associations. This aligns with prior evidence highlighting GrimAge – particularly in its updated v2 implementation – as a sensitive marker of health-related aging [36,55,56]. PhenoAge, by contrast, showed only few nominal associations in our analyses. This is somewhat in contrast to previous work which has linked PhenoAge to cognitive or functional outcomes [57], suggesting it can capture aspects of aging relevant to cognitive decline. However, and in line with our results, these prior associations appeared weaker and less consistent than those observed for GrimAge [57]. Likewise, we only observed a few nominal association signals with fourth-generation “causal” clocks recently proposed by Ying et al. [16]. Interestingly, inter-clock correlation patterns between YingDamAge and YingAdaptAge differed from those reported in the original publication. In BASE-II, they correlate strongly (R = 0.9), whereas the original publication refers to near independence among all three measures (R = 0.14). This may be a reflection of the older age structure of our sample, as these clocks were reported to be less stable in older individuals [15]. Still, the lack of a significant association with frailty – despite its inclusion in the original training framework – was unexpected and may suggest limited generalizability of the Ying clocks to older cohorts.

In contrast to the abovementioned clocks, DunedinPACE (the only third-generation DNAm clock analyzed here) stood out most clearly in essentially all our analyses. It was the only clock to show robust, consistent, and comparatively strong associations with both cognitive performance paradigms (cross-sectional and longitudinal), as well as frailty. This aligns well with findings reported by the developers of DunedinPACE and others, who have shown that faster biological aging rates estimated by this clock predict lower cognitive functioning, poorer processing speed, and steeper cognitive decline [14,22,52]. In several aspects our study is similar (both conceptually and in results) to the work by Savin et al. [22] who have investigated the association between three DNAm clocks (DunedinPACE, PhenoAge and GrimAge) and global cognition in the Framingham Offspring Cohort. They, too, found a strong link between DunedinPACE and cognitive decline [22]. Our work extends these prior analyses by studying a larger number of cognitive tests, a more diverse array of DNAm clocks (including fourth-(causal) and fifth-(system-specific) generation clocks, which were not available when Savin et al. performed their analyses) and investigating two DNAm measurement time points.

A major novel finding is the observation of strong and consistent associations of the fifth-generation SystemsAge framework with cognitive performance (and frailty). Our results independently replicate the original results by Sehgal et al., confirming robust cross-sectional associations, in particular for the Inflammation and general SystemsAge clocks. The prominent effect of the Inflammation clock provides further epigenetic evidence for the prominent role of ’inflammaging’ in cognitive decline [58]. Crucially, by utilizing a comprehensive cognitive test battery rather than broad clinical screening tools, we substantially extend these foundational findings by demonstrating that the general, Inflammation and Hormone SystemsAge clocks also show robust longitudinal associations with cognitive trajectories across specific performance domains. In contrast, we could not replicate the previously reported effects for the Brain SystemsAge clock, which likely reflects our focus on subtle functional changes rather than broad cognitive impairment.

With respect to the associated cognitive outcomes, we note a preponderance towards tests covering the Gf (general fluid intelligence) and WM (working memory) domains across all analysis paradigms. Furthermore, DSST performance also ranked very high in the list of associated cognitive tests. In contrast, episodic memory (EM) performance was not strongly associated. This dissociation aligns with the neurobiological architecture of these domains. Gf, WM, and the DSST are inherently constrained by processing speed and global white matter connectivity[59,60], possibly making them vulnerable to the diffuse, systemic biological aging process captured by peripheral DNAm clocks. Conversely, EM relies on more localized medial temporal networks and often benefits from semantic scaffolding, which may temporarily buffer it against subclinical systemic decay [61]. Consequently, blood-based epigenetic aging appears to primarily index the early erosion of broad neural network efficiency rather than the degradation of localized memory circuits. Key strengths of our study include the use of a well-characterized and comparatively large longitudinal cohort of elderly individuals, a very comprehensive cognitive test battery ascertained at multiple time points, and the utilization of DNAm clocks from all development generations, including those from the SystemsAge framework which were only published six months ago at the time of writing. Notwithstanding these strengths, we note the following potential limitations that should be considered when interpreting our results. First, all types of cognitive test scores have limited measurement precision. Day-to-day fluctuations – often unrelated to stable cognitive ability – introduce noise and complicate the differentiation between true cognitive change and short-term variability (Salthouse, 2007). Second, our study relies on DNAm measured in peripheral blood. While the use of easily accessible tissues is standard practice for developing (epigenetic) biomarkers, it traditionally limits direct conclusions about brain-specific mechanisms due to its merely partial correlation with DNAm patterns in the brain [62]. To a degree, we addressed this limitation by incorporating the SystemsAge framework, which is specifically designed to proxy organ- and system-specific aging directly from blood methylation profiles. Third, the BASE-II dataset consists exclusively of older, predominantly Northern European adults from the Berlin metropolitan area. While this homogeneity reduces environmental and sociodemographic confounding, it limits generalizability to younger individuals or other ancestries. Fourth, although the effective sample size of ∼1,000 individuals is comparatively large in the context of epigenomic projects, it remains modest relative to typical complex-trait genome-wide association studies (GWAS) [63]. While the use of quantitative traits likely mitigated some loss of statistical power [64], subtle associations may still have gone undetected.

## 6 Conclusions

In summary, our study provides comprehensive evidence that DNAm-based measures of biological aging differ substantially in their ability to capture cognitive functioning, both cross-sectionally and longitudinally. Using a broad range of current DNAm clock algorithms, DunedinPACE showed the strongest and most consistent associations with cognitive performance, followed by the newly introduced SystemsAge clocks. The other tested clocks – including first-generation algorithms, a telomere-length estimator, and causally informed fourth-generation clocks – showed no noteworthy or consistent association patterns. Our findings highlight the importance of selecting appropriate biological aging measures in studies of cognitive function and underscore the potential of DunedinPACE and system-specific DNAm clocks for improving the early detection of cognitive aging.

## Data Availability

Summary statistics for all analyses have been made available in the Supplementary Material. Access to individual-level DNAm and cognitive data is restricted but can be shared with qualified researchers upon reasonable request from the corresponding author and after approval from the BASE-II steering committee.

## 7 Abbreviations

BASE-II: Berlin Aging Study II;
CpG: Cytosine-Guanine-Dinucleotide;
DNA: Desoxyribonucleic acid;
DNAm: DNA-methylation;
DNAm-Age: epigenetic age;
DSST: Digit Symbol Substitution Test;
EM: episodic memory;
FA: Figural Analogies;
FDR: False discovery rate;
FP: Face-Profession Task;
Gf: fluid intelligence;
IKMB: Institute for clinical molecular biology;
LU: Letter Updating Task;
MPIHD: Max-Planck-Institute for human development;
NNB: Number-N-Back Task;
OL: Object Location Task;
PC: Principal Component;
PCA: Principal Component Analysis;
PP: Practical Problem Task;
QC: Quality control;
SD: standard deviation;
SNP: Single nucleotide polymorphism;
SU: Spatial Updating Task;
VLMT: Verbal Learning and Memory Test;
WHO: World Health Organization;
WM: working memory

## 8 Declarations

### 8.1 Ethics approval and consent to participate

The BASE-II/GendAge studies involving human participants were approved by the ethics committee of the Charité-Universitätsmedizin Berlin (approval numbers: EA2/144/16, EA2/029/09) and the University of Lübeck (approval numbers AZ19-390A and 19-391A). GendAge is registered in the German Clinical Trials Register (Study-ID: DRKS00016157). The cognitive tests were approved by the Ethics Committee of the Max Planck Institute for Human Development, Berlin (approval number: LIP-2012-04). The participants provided their written informed consent to participate in this study.

### 8.2 Consent for publication

Not applicable

### 8.6 Authors’ contributions

LB: Writing – original draft, Conceptualization, Supervision, Funding acquisition, Project administration. AMW: Writing – original draft, Conceptualization, Methodology, Formal analysis, Visualization. MPJ: Writing – original draft, Methodology, Formal analysis, Visualization. JH: Methodology. VD: Data generation. VMV: Methodology. UL: Funding acquisition, Data generation. CML: Funding acquisition, Data generation, Supervision. ID: Methodology, Data curation, Funding acquisition. SD: Data curation, Investigation, Methodology

## 8.7 Acknowledgements

We are deeply indebted to all individuals who have agreed to participate in BASE-II and the follow-up GendAge assessments. We would further like to thank all staff members and students involved in the medical and cognitive examinations of participants and documentation. We further thank Tanja Wesse and colleagues at IKMB for generating the DNAm data.

We acknowledge the high-performance environment (“OmicsCluster”) at the University of Lübeck where most data processing and analysis steps of this study were run.

## 8.8 Generative AI statement

This manuscript is based on data from a M.Sc. thesis (in German) of Anna Marit Weißenburg, successfully defended on Sept 11, 2025 before the MINT faculty of University of Lübeck. ChatGPT (OpenAI (2025) [Version o4]) and Gemini 3 (Google (2026)) were used to adapt the text of this M.Sc. thesis into the initial version of the current manuscript, which was subsequently substantially altered by (human) co-authors.

## References

1. World Health Organization. Progress report on the United Nations Decade of Healthy Ageing, 2021-2023 [Internet]. 2023. Licence: CC BY-NC-SA 3.0 IGO

2. Gianfredi V, Nucci D, Pennisi F, Maggi S, Veronese N, Soysal P. Aging, longevity, and healthy aging: the public health approach. Aging Clin Exp Res. 2025;37:125. 10.1007/s40520-025-03021-8

3. Margiotti K, Monaco F, Fabiani M, Mesoraca A, Giorlandino C. Epigenetic Clocks: In Aging-Related and Complex Diseases. Cytogenet Genome Res. 2023;163:247–56. 10.1159/000534561

4. Teschendorff AE, Horvath S. Epigenetic ageing clocks: statistical methods and emerging computational challenges. Nat Rev Genet. 2025;26:350–68. 10.1038/s41576-024-00807-w

5. Jaenisch R, Bird A. Epigenetic regulation of gene expression: how the genome integrates intrinsic and environmental signals. Nat Genet. 2003;33:245–54. 10.1038/ng1089

6. Moore LD, Le T, Fan G. DNA Methylation and Its Basic Function. Neuropsychopharmacol. 2013;38:23–38. 10.1038/npp.2012.112

7. Nabais MF, Gadd DA, Hannon E, Mill J, McRae AF, Wray NR. An overview of DNA methylation-derived trait score methods and applications. Genome Biol. 2023;24:28. 10.1186/s13059-023-02855-7

8. Kornicka K, Marycz K, Marędziak M, Tomaszewski KA, Nicpoń J. The effects of the DNA methyltranfserases inhibitor 5-Azacitidine on ageing, oxidative stress and DNA methylation of adipose derived stem cells. J Cellular Molecular Medi. 2017;21:387–401. 10.1111/jcmm.12972

9. Johnson ND, Conneely KN. The role of DNA methylation and hydroxymethylation in immunosenescence. Ageing Research Reviews. 2019;51:11–23. 10.1016/j.arr.2019.01.011

10. Hannum G, Guinney J, Zhao L, Zhang L, Hughes G, Sadda S, et al. Genome-wide Methylation Profiles Reveal Quantitative Views of Human Aging Rates. Molecular Cell. 2013;49:359–67. 10.1016/j.molcel.2012.10.016

11. Horvath S. DNA methylation age of human tissues and cell types. Genome Biol. 2013;14:3156. 10.1186/gb-2013-14-10-r115

12. Levine ME, Lu AT, Quach A, Chen BH, Assimes TL, Bandinelli S, et al. An epigenetic biomarker of aging for lifespan and healthspan. Aging. 2018;10:573–91. 10.18632/aging.101414

13. Lu AT, Quach A, Wilson JG, Reiner AP, Aviv A, Raj K, et al. DNA methylation GrimAge strongly predicts lifespan and healthspan. Aging. 2019;11:303–27. 10.18632/aging.101684

14. Belsky DW, Caspi A, Corcoran DL, Sugden K, Poulton R, Arseneault L, et al. DunedinPACE, a DNA methylation biomarker of the pace of aging. eLife. 2022;11:e73420. 10.7554/eLife.73420

15. Ying K, Liu H, Tarkhov AE, Sadler MC, Lu AT, Moqri M, et al. Causality-enriched epigenetic age uncouples damage and adaptation. Nat Aging. 2024;4:231–46. 10.1038/s43587-023-00557-0

16. Ying K, Liu H, Tarkhov AE, Sadler MC, Lu AT, Moqri M, et al. Causality-Enriched Epigenetic Age Uncouples Damage and Adaptation. Nat Aging. 2024;4:231–46. 10.1038/s43587-023-00557-0

17. Sehgal R, Markov Y, Qin C, Meer M, Hadley C, Shadyab AH, et al. Systems Age: a single blood methylation test to quantify aging heterogeneity across 11 physiological systems. Nat Aging. Nature Publishing Group; 2025;5:1880–96. 10.1038/s43587-025-00958-3

18. Grodstein F, Lemos B, Yu L, Klein H-U, Iatrou A, Buchman AS, et al. The association of epigenetic clocks in brain tissue with brain pathologies and common aging phenotypes. Neurobiology of Disease. 2021;157:105428. 10.1016/j.nbd.2021.105428

19. Higgins-Chen AT, Thrush KL, Levine ME. Aging biomarkers and the brain. Seminars in Cell & Developmental Biology. 2021;116:180–93. 10.1016/j.semcdb.2021.01.003

20. Degerman S, Josefsson M, Nordin Adolfsson A, Wennstedt S, Landfors M, Haider Z, et al. Maintained memory in aging is associated with young epigenetic age. Neurobiology of Aging. Elsevier BV; 2017;55:167–71. 10.1016/j.neurobiolaging.2017.02.009

21. Beydoun MA, Shaked D, Tajuddin SM, Weiss J, Evans MK, Zonderman AB. Accelerated epigenetic age and cognitive decline among urban-dwelling adults. Neurology. Ovid Technologies (Wolters Kluwer Health); 2020;94:e613–25. 10.1212/wnl.0000000000008756

22. Savin MJ, Wang H, Pei H, Aiello AE, Assuras S, Caspi A, et al. Association of a pace of aging epigenetic clock with rate of cognitive decline in the Framingham Heart Study Offspring Cohort. Alz & Dem Diag Ass & Dis Mo [Internet]. Wiley; 2024 [cited 2025 July 22];16. 10.1002/dad2.70038

23. Vetter VM, Drewelies J, Homann J, Düzel S, Deecke L, Jawinski P, et al. Comprehensive Comparison of Sixteen Markers of Biological Aging: Cross-Sectional and Longitudinal Results from the Berlin Aging Study II (BASE-II) [Internet]. medRxiv; 2025 [cited 2026 Mar 16]. p. 2025.04.09.25325514. 10.1101/2025.04.09.25325514

24. Sibbett RA, Altschul DM, Marioni RE, Deary IJ, Starr JM, Russ TC. DNA methylation-based measures of accelerated biological ageing and the risk of dementia in the oldest-old: a study of the Lothian Birth Cohort 1921. BMC Psychiatry. 2020;20:91. 10.1186/s12888-020-2469-9

25. Bertram L, Böckenhoff A, Demuth I, Düzel S, Eckardt R, Li S-C, et al. Cohort profile: The Berlin Aging Study II (BASE-II). Int J Epidemiol. 2014;43:703–12. 10.1093/ije/dyt018

26. Düzel S, Voelkle MC, Düzel E, Gerstorf D, Drewelies J, Steinhagen-Thiessen E, et al. The Subjective Health Horizon Questionnaire (SHH-Q): Assessing Future Time Perspectives for Facets of an Active Lifestyle. Gerontology. 2016;62:345–53. 10.1159/000441493

27. Demuth I, Banszerus V, Drewelies J, Düzel S, Seeland U, Spira D, et al. Cohort profile: follow-up of a Berlin Aging Study II (BASE-II) subsample as part of the GendAge study. BMJ Open. 2021;11:e045576. 10.1136/bmjopen-2020-045576

28. Wang Y, Hannon E, Grant OA, Gorrie-Stone TJ, Kumari M, Mill J, et al. DNA methylation-based sex classifier to predict sex and identify sex chromosome aneuploidy. BMC Genomics. 2021;22:484. 10.1186/s12864-021-07675-2

29. Sommerer Y, Dobricic V, Schilling M, Ohlei O, Bartrés-Faz D, Cattaneo G, et al. Epigenome-Wide Association Study in Peripheral Tissues Highlights DNA Methylation Profiles Associated with Episodic Memory Performance in Humans. Biomedicines. 2022;10:2798. 10.3390/biomedicines10112798

30. Sommerer Y, Dobricic V, Schilling M, Ohlei O, Sabet SS, Wesse T, et al. Entorhinal cortex epigenome-wide association study highlights four novel loci showing differential methylation in Alzheimer’s disease. Alzheimers Res Ther. 2023;15:92. 10.1186/s13195-023-01232-7

31. Du P, Zhang X, Huang C-C, Jafari N, Kibbe WA, Hou L, et al. Comparison of Beta-value and M-value methods for quantifying methylation levels by microarray analysis. BMC Bioinformatics. Springer Science and Business Media LLC; 2010;11:587. 10.1186/1471-2105-11-587

32. Gorrie-Stone TJ, Smart MC, Saffari A, Malki K, Hannon E, Burrage J, et al. Bigmelon: tools for analysing large DNA methylation datasets. Bioinformatics. 2019;35:981–6. 10.1093/bioinformatics/bty713

33. Zhou W, Laird PW, Shen H. Comprehensive characterization, annotation and innovative use of Infinium DNA methylation BeadChip probes. Nucleic acids research. Oxford University Press; 2017;45:e22–e22.

34. Nordlund J, Bäcklin CL, Wahlberg P, Busche S, Berglund EC, Eloranta M-L, et al. Genome-wide signatures of differential DNA methylation in pediatric acute lymphoblastic leukemia. Genome biology. Springer; 2013;14:1–15.

35. Sørensen Ø, Fjell AM, Walhovd KB. Longitudinal Modeling of Age-Dependent Latent Traits with Generalized Additive Latent and Mixed Models. Psychometrika. 2023;88:456–86. 10.1007/s11336-023-09910-z

36. Lu AT, Binder AM, Zhang J, Yan Q, Reiner AP, Cox SR, et al. DNA methylation GrimAge version 2. Aging. 2022;14:9484–549. 10.18632/aging.204434

37. Ye Q, Apsley AT, Etzel L, Hastings WJ, Kozlosky JT, Walker C, et al. Telomere length and chronological age across the human lifespan: A systematic review and meta-analysis of 414 study samples including 743,019 individuals. Ageing Research Reviews. 2023;90:102031. 10.1016/j.arr.2023.102031

38. Lu AT, Seeboth A, Tsai P-C, Sun D, Quach A, Reiner AP, et al. DNA methylation-based estimator of telomere length. Aging. 2019;11:5895–923. 10.18632/aging.102173

39. Ying K, Paulson S, Eames A, Tyshkovskiy A, Li S, Eynon N, et al. A unified framework for systematic curation and evaluation of aging biomarkers. Nat Aging. 2025;5:2323–39. 10.1038/s43587-025-00987-y

40. Thrush KL, Higgins-Chen AT, Liu Z, Levine ME. R methylCIPHER: A Methylation Clock Investigational Package for Hypothesis-Driven Evaluation & Research [Internet]. bioRxiv; 2022 [cited 2026 Mar 11]. p. 2022.07.13.499978. 10.1101/2022.07.13.499978

41. DNAm Clock Foundation. DNAm Clock Foundation Calculator [Internet]. DNAm Clock Foundation. 2023. https://dnamage.clockfoundation.org/

42. Benjamini Y, Hochberg Y. Controlling the False Discovery Rate: A Practical and Powerful Approach to Multiple Testing. Royal Statistical Society Journal Series B: Methodological. 1995;57:289–300. 10.1111/j.2517-6161.1995.tb02031.x

43. Gale CR, Marioni RE, Harris SE, Starr JM, Deary IJ. DNA methylation and the epigenetic clock in relation to physical frailty in older people: the Lothian Birth Cohort 1936. Clinical Epigenetics. 2018;10:101. 10.1186/s13148-018-0538-4

44. Verschoor CP, Lin DTS, Kobor MS, Mian O, Ma J, Pare G, et al. Epigenetic age is associated with baseline and 3-year change in frailty in the Canadian Longitudinal Study on Aging. Clin Epigenetics. 2021;13:163. 10.1186/s13148-021-01150-1

45. Vetter VM, Kalies CH, Sommerer Y, Spira D, Drewelies J, Regitz-Zagrosek V, et al. Relationship Between 5 Epigenetic Clocks, Telomere Length, and Functional Capacity Assessed in Older Adults: Cross-Sectional and Longitudinal Analyses. J Gerontol A Biol Sci Med Sci. 2022;77:1724–33. 10.1093/gerona/glab381

46. Fried LP, Tangen CM, Walston J, Newman AB, Hirsch C, Gottdiener J, et al. Frailty in Older Adults: Evidence for a Phenotype. The Journals of Gerontology Series A: Biological Sciences and Medical Sciences. 2001;56:M146–57. 10.1093/gerona/56.3.M146

47. Buchmann N, Spira D, König M, Demuth I, Steinhagen-Thiessen E. Frailty and the Metabolic Syndrome — Results of the Berlin Aging Study II (BASE-II). The Journal of Frailty & Aging. 2019;8:169–75. 10.14283/jfa.2019.15

48. Govender P, Ghai M, Okpeku M. Sex-specific DNA methylation: impact on human health and development. Mol Genet Genomics. Springer Science and Business Media LLC; 2022;297:1451–66. 10.1007/s00438-022-01935-w

49. Tesfaye M, Spindola LM, Stavrum A-K, Shadrin A, Melle I, Andreassen OA, et al. Sex effects on DNA methylation affect discovery in epigenome-wide association study of schizophrenia. Mol Psychiatry. Springer Science and Business Media LLC; 2024;29:2467–77. 10.1038/s41380-024-02513-9

50. Horn JL, Cattell RB. Age differences in fluid and crystallized intelligence. Acta Psychol (Amst). 1967;26:107–29. 10.1016/0001-6918(67)90011-x

51. Mitchell DJ, Mousley ALS, Shafto MA, Duncan J. Neural Contributions to Reduced Fluid Intelligence across the Adult Lifespan. J Neurosci. 2023;43:293–307. 10.1523/JNEUROSCI.0148-22.2022

52. Nguyen S, McEvoy LK, Espeland MA, Whitsel EA, Lu A, Horvath S, et al. Associations of Epigenetic Age Estimators With Cognitive Function Trajectories in the Women’s Health Initiative Memory Study. Neurology. 2024;103:e209534. 10.1212/WNL.0000000000209534

53. Marioni RE, Shah S, McRae AF, Ritchie SJ, Muniz-Terrera G, Harris SE, et al. The epigenetic clock is correlated with physical and cognitive fitness in the Lothian Birth Cohort 1936. Int J Epidemiol. 2015;44:1388–96. 10.1093/ije/dyu277

54. Bressler J, Marioni RE, Walker RM, Xia R, Gottesman RF, Windham BG, et al. Epigenetic Age Acceleration and Cognitive Function in African American Adults in Midlife: The Atherosclerosis Risk in Communities Study. J Gerontol A Biol Sci Med Sci. 2020;75:473–80. 10.1093/gerona/glz245

55. Maddock J, Castillo-Fernandez J, Wong A, Cooper R, Richards M, Ong KK, et al. DNA Methylation Age and Physical and Cognitive Aging. J Gerontol A Biol Sci Med Sci. 2019;75:504–11. 10.1093/gerona/glz246

56. Zheng Y, Habes M, Gonzales M, Pomponio R, Nasrallah I, Khan S, et al. Mid-life epigenetic age, neuroimaging brain age, and cognitive function: coronary artery risk development in young adults (CARDIA) study. Aging (Albany NY). 2022;14:1691–712. 10.18632/aging.203918

57. McCrory C, Fiorito G, Hernandez B, Polidoro S, O’Halloran AM, Hever A, et al. GrimAge Outperforms Other Epigenetic Clocks in the Prediction of Age-Related Clinical Phenotypes and All-Cause Mortality. J Gerontol A Biol Sci Med Sci. 2020;76:741–9. 10.1093/gerona/glaa286

58. Soraci L, Corsonello A, Paparazzo E, Montesanto A, Piacenza F, Olivieri F, et al. Neuroinflammaging: A Tight Line Between Normal Aging and Age-Related Neurodegenerative Disorders. Aging Dis. 2024;15:1726–47. 10.14336/AD.2023.1001

59. Salthouse TA. The processing-speed theory of adult age differences in cognition. Psychol Rev. 1996;103:403–28. 10.1037/0033-295x.103.3.403

60. Madden DJ, Bennett IJ, Song AW. Cerebral white matter integrity and cognitive aging: contributions from diffusion tensor imaging. Neuropsychol Rev. 2009;19:415–35. 10.1007/s11065-009-9113-2

61. Park DC, Reuter-Lorenz P. The Adaptive Brain: Aging and Neurocognitive Scaffolding. Annu Rev Psychol. 2009;60:173–96. 10.1146/annurev.psych.59.103006.093656

62. Sommerer Y, Ohlei O, Dobricic V, Oakley DH, Wesse T, Sedghpour Sabet S, et al. A correlation map of genome-wide DNA methylation patterns between paired human brain and buccal samples. Clin Epigenet. 2022;14:139. 10.1186/s13148-022-01357-w

63. Wu X-R, Wu B-S, Kang J-J, Chen L-M, Deng Y-T, Chen S-D, et al. Contribution of copy number variations to education, socioeconomic status and cognition from a genome-wide study of 305,401 subjects. Mol Psychiatry. Springer Science and Business Media LLC; 2025;30:889–98. 10.1038/s41380-024-02717-z

64. Yang J, Wray NR, Visscher PM. Comparing apples and oranges: equating the power of case-control and quantitative trait association studies. Genet Epidemiol. 2010;34:254–7. 10.1002/gepi.20456

