## Supplementary Material for "Biological age acceleration measured by DunedinPACE associates most consistently with cognitive decline in elderly individuals"

### 1. Supplementary Methods

#### 1.1 Measurements of cognitive phenotypes

*Digit Symbol Substitution Test (DSST):* The Digit Symbol Substitution Test is associated with perceptual speed and other cognitive domains. Participants were shown a code box containing nine digit-symbol pairs, along with a sequence of digits. They were instructed to draw the corresponding symbols from the code box underneath the digits within 90 seconds. The test is part of the Wechsler Adult Intelligence Scale-Revised [1].

*Figural Analogies (FA):* Figural Analogies serve as an indicator of logical reasoning and are associated with visuospatial reasoning and fluid intelligence (Gf) [2]. Participants were presented with visual analogy items following the pattern "A is to B as C is to ?". A pair of figures (A and B) and a target figure (C) were given. Participants selected the figure that best completed the analogy from five possible options. After a sample item, a series of test items increasing in difficulty was administered. The test ended after three consecutive errors, reaching the final item, or a time limit of 10 minutes.

*Face-Profession Task (FP):* The Face-Profession Task is based on the 1-to-1 source-fact paradigm described by Schacter et al. (1994) and assesses associative memory (a component of episodic memory) [3]. Participants viewed 45 face-profession pairs for 3.5 seconds each and judged whether the pairings were a good match. After a 3-minute delay, 54 face-profession pairings were presented, including 27 previously seen pairs, 9 novel pairs, and 18 recombined pairs (familiar faces with new professions). Participants indicated whether they had seen each pair before and rated their confidence.

*Letter Updating Task (LU):* The Letter Updating Task measures working memory. Participants viewed sequences of 7, 9, 11, or 13 letters and, after each sequence, had to select the last three letters in correct order from four response options [4].

*Number-N-Back Task (NNB):* The Number-N-Back Task (also referred to as the 3-back numerical task [5]) assesses working memory. Participants saw a sequence of three digits followed by a stream of 30 sequences, each consisting of three digits. For each sequence, they had to indicate whether it matched the sequence presented three trials earlier. The task included four practice rounds and six test rounds.

*Object Location Task (OL):* The Object Location Task is part of the web-based training program by Schmiedek et al. and assesses spatial and episodic memory [5]. Participants viewed a sequence of 12 colored images presented at different locations in a 6×6 grid. Afterwards, they were asked to drag each image from the screen edge to its correct location using the mouse. One practice and two test trials were completed.

*Practical Problem Task (PP):* The Practical Problem Task, a measure of fluid intelligence, presents 12 everyday problems such as interpreting a bus schedule, medication instructions, or street maps [6]. Each item appeared at the top of the screen along with five response options. After one practice problem, participants completed problems in order of increasing difficulty until either three consecutive errors occurred, the time limit of 10 minutes was reached, or the last problem was completed.

*Spatial Updating Task (SU):* The Spatial Updating Task assesses working memory and is based on the Memory Updating Spatial Task by Schmiedek et al. [5]. Participants viewed two or three 3×3 grids for four seconds, each containing a blue dot at one location. They then had to update the position of the dot based on directional arrows shown for 2.5 seconds below each grid, with six updating steps shown at 0.5-second intervals. Participants clicked on the final position of the dot. After ten practice trials, ten test trials followed.

*Verbal Learning and Memory Test (VLMT):* The Verbal Learning and Memory Test (also referred to as the Rey Auditory Verbal Learning Test, RAVLT) measures episodic memory and was developed by Rey (1964) and adapted into German by Helmstaedter & Durwen (1990) [7]. The test consists of five learning trials during which a list of 15 words is read aloud and recalled by participants. A second list of 15 words is presented and recalled once. Immediately afterward, and again after a 30-minute delay, participants recalled the first list. At T0, delayed recall was also assessed after seven days.

*Latent Cognitive Factors: Episodic Memory (EM), Working Memory (WM), Fluid Intelligence (Gf):* To derive latent cognitive factors, a confirmatory factor analysis based on a three-factor model was conducted (for more information see Düzel et al., 20216) [8]. Correlations between latent factors and individual cognitive tests are shown in Supplementary Figure S1.

*Episodic memory (EM)* comprised the Scene Encoding (not used in individual analyses), Verbal Learning and Memory Test, Face-Profession Task, and Object Location Task.

*Working memory (WM)* was derived from the Spatial Updating Task, Letter Updating Task, and Number-N-Back Task.

*Fluid intelligence (Gf)* was defined by the Practical Problem Task, Figural Analogies, and Letter Series (not used in individual analyses).

*Outlier removal of cognitive test results:* For all cognitive abilities, outliers that deviated by four standard deviations from the mean of the respective test were excluded. Some individuals were additionally removed due to uncertain data with values of 0 in FA, SU, WM, EM and Gf, values above 22 for FA and an outlier with a value of 58 in LU (T2). With the SexEstimation on Biolearn one individual with Turner syndrome and one with Klinefelter syndrome were identified and excluded.

### 2. Supplementary Results

#### 2.1 Comparison between age estimates from Biolearn and the DNAmAge Calculator

Epigenetic age estimation has traditionally been performed using the DNAmAge Calculator website (URL: https://dnamage.clockfoundation.org/) or individual published scripts in R or Python. The open-source library Biolearn consolidates various epigenetic clocks into a single, standardized Python package [9]. One limitation of the DNAmAge Calculator is the requirement to upload DNAm data along with participants' age and sex; for data privacy reasons, locally running scripts are preferable. Biolearn allows for local computation of a wide range of epigenetic clocks and other models, offering greater flexibility than the DNAmAge Calculator. Given that Biolearn represents a pre-publication version and prior work in our group has been based on the DNAmAge-Calculator, we assessed the comparability between both tools regarding the prediction of biological age from DNAm data. Specifically, we examined the Horvath (v1) [10], Hannum [11], PhenoAge [12], GrimAge (v2) [13] clocks, DNAmTL (telomere length) [14], and sex prediction [15], as the DNAmAge-Calculator does not provide outputs for all available clocks in Biolearn.

For the Horvath (v1) clock, results from both tools showed non-systematic deviations ranging from -3.14 to 2.65 years. According to the DNAmAge-Calculator documentation, this discrepancy may be attributed to a normalization step aligning the input data to the original training set. When this normalization was disabled (in the older version of the calculator – DNAmAge-Calculators, UCLA Horvath Lab, 2013), the discrepancy disappeared, suggesting that the variation was largely due to this preprocessing step. Since the details of this normalization are not fully disclosed and may impair comparability across clocks, we chose to use Biolearn-derived estimates for the Horvath (v1) clock in all main analyses. However, to ensure robustness, cognitive associations were also tested using the normalized DNAmAge-Calculator predictions. The single nominally significant association observed with the DNAmAge-Calculator was not nominally significant with Biolearn. However, both methods yielded consistent directionality and highly similar effect sizes (Supplementary Table S11). This consistency supports the validity of the Biolearn predictions. For better comparability with other epigenetic clocks, results derived from Biolearn are preferable. Given that associations between Horvath (v1) age acceleration and cognitive performance showed only one nominally significant and no FDR-significant findings, minor differences between implementations can be considered negligible. However, in analyses where Horvath (v1) age predictions show stronger significance, it may be worthwhile to revisit the comparison between both implementations.

Regarding the Hannum clock, Biolearn consistently predicted older ages, with a systematic offset of +10.25 years compared to the DNAmAge-Calculator. In contrast, second-generation clocks PhenoAge and GrimAge (v2) showed minimal systematic differences between the tools. The systematic discrepancy in Hannum age estimates arises due to the absence of six CpGs in the methylation dataset used here, leading to different imputation approaches. In Biolearn, all clocks except DunedinPACE and the SexEstimation model were imputed using a hybrid method based on the SeSAMe 450k gold standard [9]. The DNAmAge Calculator does not provide detailed documentation on its imputation strategy [17]. Systematic differences in Hannum, PhenoAge, and GrimAge (v2) clocks can be disregarded since the association analyses were conducted using age acceleration residuals, which eliminate such systematic biases.

Predictions of telomere length also varied substantially between tools. DNAmTL estimates from Biolearn were on average 15.85 kb higher than those from the DNAmAge-Calculator. The systematic discrepancy in telomere length predictions results from the transformation and definition of the DNAmTL output in Biolearn (version 0.6.5). This issue can be resolved by analyzing DNAmTLadj values.

Lastly, sex prediction accuracy was high with both methods, but Biolearn's *SexEstimation* identified two participants with atypical karyotypes not highlighted by the DNAmAge Calculator: one individual with Klinefelter syndrome (47, XXY), previously documented, and one previously undiagnosed case likely consistent with Turner syndrome (45, X0). The latter was supported by clinical indicators including short stature (165 cm), undetectable estrogen levels, and nulliparity. The improved accuracy of sex prediction in Biolearn is likely due to the substantially larger number of CpGs used for sex estimation compared to the DNAmAge Calculator.

Overall, the results from Biolearn and the DNAmAge Calculator are largely comparable. However, due to enhanced data privacy and the inclusion of additional epigenetic clocks, Biolearn represents a preferable approach for future epigenetic age estimations.

### 3. Supplementary Figures

**Supplementary Figure S1: Pearson-Correlation of all cognition tests with their correlation-coefficient.**

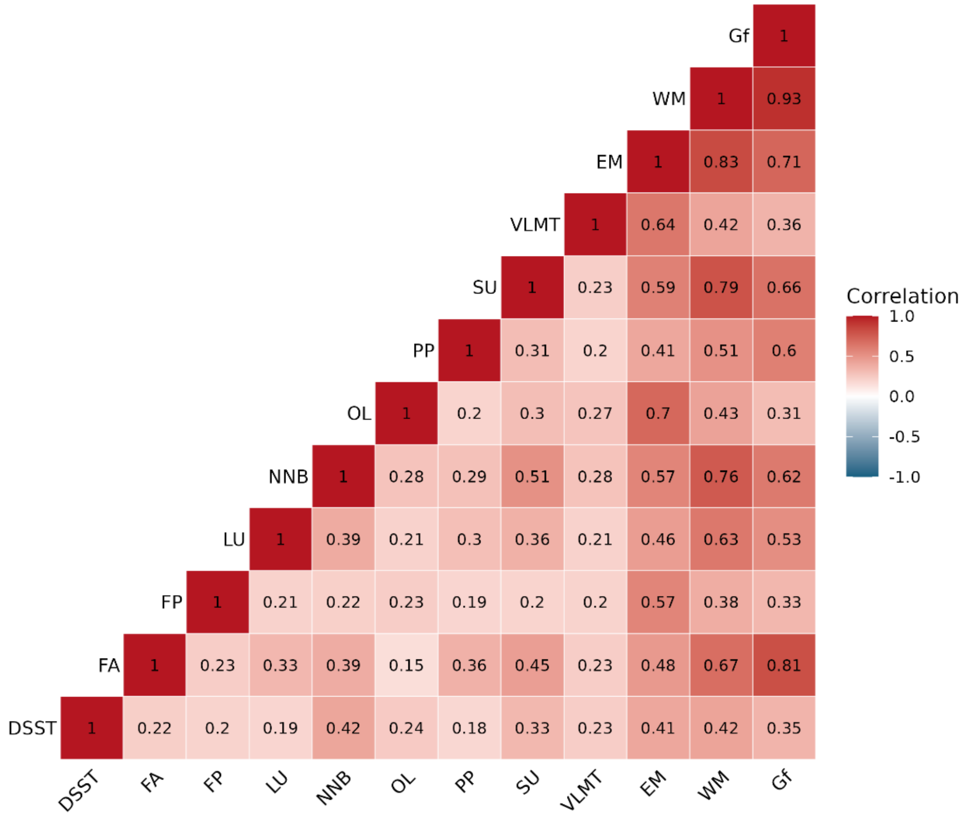


Legend: The cognition tests were z-standardized before analysis. For brevity, cognitive tests are denoted by their abbreviations. Full names are provided in the list of abbreviations and the Methods section.

### 4. References


1. Wechsler D. WAIS-R Manual: Wechsler Adult Intelligence Scale-revised [Internet]. Psychological Corporation; 1981. https://books.google.de/books?id=7IMiNgAACAAJ

2. Lindenberger U, Mayr U, Kliegl R. Speed and intelligence in old age. Psychology and Aging. 1993;8:207–20. https://doi.org/10.1037/0882-7974.8.2.207

3. Schacter DL, Osowiecki D, Kaszniak AW, Kihlstrom JF, Valdiserri M. Source memory: Extending the boundaries of age-related deficits. Psychology and Aging. 1994;9:81–9. https://doi.org/10.1037/0882-7974.9.1.81

4. Artuso C, Palladino P. How sublexical association strength modulates updating: Cognitive and strategic effects. Mem Cogn. 2018;46:285–97. https://doi.org/10.3758/s13421-017-0764-6

5. Schmiedek F, Lövdén M, Lindenberger U. Hundred Days of Cognitive Training Enhance Broad Cognitive Abilities in Adulthood: Findings from the COGITO Study. Front Aging Neurosci [Internet]. Frontiers; 2010 [cited 2026 Mar 23];2. https://doi.org/10.3389/fnagi.2010.00027

6. Lindenberger U, Mayr U, Kliegl R. Speed and intelligence in old age. Psychology and Aging. US: American Psychological Association; 1993;8:207–20. https://doi.org/10.1037/0882-7974.8.2.207

7. Helmstaedter C, Durwen HF. VLMT: Verbaler Lern- und Merkfähigkeitstest: Ein praktikables und differenziertes Instrumentarium zur Prüfung der verbalen Gedächtnisleistungen. [VLMT: A useful tool to assess and differentiate verbal memory performance.]. Schweizer Archiv für Neurologie, Neurochirurgie und Psychiatrie. Switzerland: Schwabe & Co; 1990;141:21–30.

8. Düzel S, Voelkle MC, Düzel E, Gerstorf D, Drewelies J, Steinhagen-Thiessen E, et al. The Subjective Health Horizon Questionnaire (SHH-Q): Assessing Future Time Perspectives for Facets of an Active Lifestyle. Gerontology. 2016;62:345–53. https://doi.org/10.1159/000441493

9. Ying K, Paulson S, Eames A, Tyshkovskiy A, Li S, Eynon N, et al. A unified framework for systematic curation and evaluation of aging biomarkers. Nat Aging. 2025;5:2323–39. https://doi.org/10.1038/s43587-025-00987-y

10. Horvath S. DNA methylation age of human tissues and cell types. Genome Biol. 2013;14:3156. https://doi.org/10.1186/gb-2013-14-10-r115

11. Hannum G, Guinney J, Zhao L, Zhang L, Hughes G, Sadda S, et al. Genome-wide Methylation Profiles Reveal Quantitative Views of Human Aging Rates. Molecular Cell. 2013;49:359–67. https://doi.org/10.1016/j.molcel.2012.10.016

12. Levine ME, Lu AT, Quach A, Chen BH, Assimes TL, Bandinelli S, et al. An epigenetic biomarker of aging for lifespan and healthspan. Aging. 2018;10:573–91. https://doi.org/10.18632/aging.101414

13. Lu AT, Binder AM, Zhang J, Yan Q, Reiner AP, Cox SR, et al. DNA methylation GrimAge version 2. Aging. 2022;14:9484–549. https://doi.org/10.18632/aging.204434

14. Lu AT, Seeboth A, Tsai P-C, Sun D, Quach A, Reiner AP, et al. DNA methylation-based estimator of telomere length. Aging. 2019;11:5895–923. https://doi.org/10.18632/aging.102173

15. Wang Y, Hannon E, Grant OA, Gorrie-Stone TJ, Kumari M, Mill J, et al. DNA methylation-based sex classifier to predict sex and identify sex chromosome aneuploidy. BMC Genomics. 2021;22:484. https://doi.org/10.1186/s12864-021-07675-2

16. Horvath Lab. DNAmAge Calculator (UCLA) [Internet]. Horvath's DNAmAge Calculator. 2013. https://dnamage.genetics.ucla.edu/

17. DNAm Clock Foundation. DNAm Clock Foundation Calculator [Internet]. DNAm Clock Foundation. 2023. https://dnamage.clockfoundation.org/
